# Predictive Modelling of COVID-19 New Cases in Algeria using An Extreme Learning Machines (ELM)

**DOI:** 10.1101/2020.09.28.20203299

**Authors:** Messaoud Djeddou, Ibrahim A. Hameed, Abolfazl Nejatian, Imed Loukam

## Abstract

In this research, an extreme learning machine (ELM) is proposed to predict the new COVID-19 cases in Algeria. In the present study, public health database from Algeria health ministry has been used to train and test the ELM models.

The input parameters for the predictive models include Cumulative Confirmed COVID-19 Cases (CCCC), Calculated COVID-19 New Cases (CCNC), and Index Day (ID).

The predictive accuracy of the seven models has been assessed via several statistical parameters. The results showed that the proposed ELM model achieved an adequate level of prediction accuracy with smallest errors (MSE= 0.16, RMSE=0.4114, and MAE= 0.2912), and highest performance’s (NSE = 0.9999, IO = 0.9988, R^2^ = 0.9999). Hence, the ELM model could be utilized as a reliable and accurate modeling approach for predicting the new COVIS-19 cases in Algeria.

The proposed ELM model, it can be used as a decision support tool to manage public health medical efforts and facilities against the COVID-19 pandemic crisis.

## 1 INTRODUCTION

Bacteria, parasites, and viruses can cause serious diseases for human health. Still, pathogenic viruses usually cause the most serious diseases and among these viruses; coronavirus is the large family of pathogenic viruses.

Infection with these types of viruses can cause respiratory, liver, gastrointestinal and neurological diseases. They are distributed among humans, birds, cattle, mice, bats and other wildlife [1–2].

World Health Organization (WHO) received notifications from Chinese authorities about many respiratory problems was linked to people who visited a local market for seafood and wildlife in Wuhan city, Hubei Province, China [3]. Virological investigation suggests that the causative agent of this pneumonia is a new coronavirus (COVID-19) [4].

This new coronavirus has made hundreds of thousands of victims around the world and becomes a global pandemic threatening human health.

Dehesh et al. [5] proposed an ARIMA model to predict the trend of confirmed cases in different countries.. They suggest that China and Thailand had almost a stable trend. The trend of South Korea was decreasing and will become stable. Iran and Italy had unstable trends. Al-qaness et al. [6] proposed an improved adaptive neuro-fuzzy inference system (ANFIS) to forecast the COVID-19 confirmed cases ten days ahead with good performances. The model was tested using two different datasets of weekly influenza confirmed cases in USA and China.

Jung et al. [7] used a statistical estimation of the confirmed case fatality risk (cCFR) and the basic reproduction number, The estimated values of the cCFR were 5.3% for Scenario 1 and 8.4% for Scenario 2. The basic reproduction number was estimated to be 2.1 and 3.2 for Scenarios 1 and 2, respectively. They argued that the current COVID-19 epidemic has a substantial potential for causing a pandemic.

Zhao et al. [8] proposed a mathematical model to estimate the real number of COVID-19 cases in the first half of January 2020. They estimated that after 17 January 2020, the cases had increased 21-fold. Nishiura et al. [9] proposed an estimation model for the infection rate of COVID-19 in Wuhan, China. They conclude that the estimated rate is 9.2%; and the death rate is 0.3% to 0.6%.

Tang et al. [10] proposed a mathematical model to estimate the transmission risk of COVID-19. They concluded that the basic reproduction number might be as high as 6.47. They also predicted the number of confirmed cases in seven days (23 to 29 January 2020).

At last, In [11], data of 47 patients were used to estimate sustained human-to-human transmission of COVID-19. The author concluded that the transmission is 0.41. However, with intense surveillance, the probability that an imported case will result in sustained transmission is only 0.012.

Since Huang el al [12] proposed the extreme learning machine (ELM), several researches based on the application of these models in various fields of science and engineering have been published to prove their significant progress over conventional neural network models.

No iterative process is required to adjust the biases and weights of the free connections between the hidden layer and the output layer, since this adjustment is adopted randomly, thus allowing a reduction in computation time. Therefore, an ELM model is remarkably efficient particularly over conventional artificial neural network to reach a global optimum, following a universal approximation of the capacity of a single-layer feed-forward network [13-14]. More other applications of the ELM model can be found in [14].

In the present research paper, the implementation of the machine learning regression model, namely extreme learning machine (ELM) is developed for the first time to predict new COVID-19 cases in Algeria.

## 2 MATERIALS AND METHODS

### 2.1 Study area

The pandemic Covid-19 in Algeria spreads from February 25, 2020. A foreign worker from Italy was tested positive for SARS-CoV-2, and then an outbreak of contagion is formed in Blida Province [15]. Sixteen members of the same family were infected with the coronavirus at a wedding party [16]. Blida Province becomes the epicenter of the coronavirus epidemic in Algeria [17].

On April 25 2020, there were 419 deaths and 3256 COVID-19 confirmed cases in Algeria.

The data used in this study include total confirmed COVID-19 cases, calculated new COVID cases, and Day index. The statistical parameters of the data used for modeling are given in Table 1.

**Table 1.** Descriptive statistics for the parameters used for New COVID-19 cases modelling

| Parameters | Min. | Max. | Mean | Std. dev. |
| --- | --- | --- | --- | --- |
| **Inputs** |  |  |  |  |
| 1- Cumulative Confirmed COVID-19 Cases (CCCC) | 3 | 3256 | 998.78 | 1063.67 |
| 2- Calculated COVID-19 New Cases (CCNC) | 0 | 185 | 57.82 | 49.56 |
| 3- Index Day (ID) | 1 | 7 | 4 | 2.01 |
| **Output** |  |  |  |  |
| **COVID-19 New Cases (CNC)** | 0 | 185 | 57.82 | 49.56 |

### 2.2. Extreme learning machines

The Extreme Learning Machine (ELM) proposed by Huang et al. [12] is a model based on a single hidden layer neural network, which after setting the weights and biases of the hidden layer, and based on a single layer feed forward network (SLFN) algorithm, allows to obtain a closed form solution for the output weights by a least squares solution. It is drawn from a continuous probability distribution function [13].

The advantages of the ELM model are [11-13]:

1- Less complex design;
2- High classification accuracy;
3- Good generalization ability;
4- Less computing time.

During training process, the weights and biases of the hidden single hidden layer feed-forward networks (SLFN) are randomly adopted and never updated. The output weights are stated using the generalized Moore-Penrose generalized inverse of the hidden layer output matrix [19].

For a set of d-dimensional vectors defined for i *i = 1, 2, …, N* training samples, the SLFN with L hidden neurons is mathematically expressed as follows [12]:

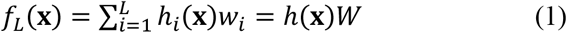

Where:

*W* = [*w*_1_, *w*_2_ … *w*_L_]^*T*^is the output weight matrix between the hidden the hidden neurons and output neurons;

*h*(***x***)= [*h*_1_, *h*_2_ … *h*_3_] is the hidden neuron outputs that represent the randomized hidden features of predictor ***x*** _i_;

*hi* (**x**)is the *i*^*th*^ hidden neuron.

The output function of the hidden neurons, *hi(***x**) can be represented as follows [14]:

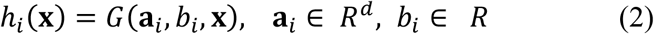

*G* (**a**_*i*_, *b*_*i*_ **x**), which is defined using hidden neuron parameters (**a**, *b)*, is a non-linear piecewise continuous function that must satisfy the ELM approximation theorem [12-13-14].

To develop the proposed ELM model, the sigmoid transfer function was adopted, which has been widely adopted in neural network-based modelling, the sigmoid equation is expressed below:

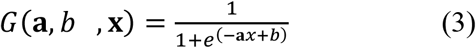

Huang et al.[14] state that the approximation error must be minimized when solving for the weights connecting the hidden and output layer (*W*) using least square fitting:

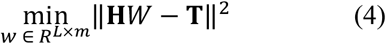

In Eq. (4), the term ‖ **H***W* − **T** ‖ is the Frobenius norm and **H** is the randomized hidden layer output matrix of the form [14]:

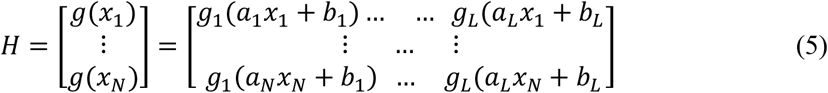

The target matrix in the data-training period is expressed in [14] as:

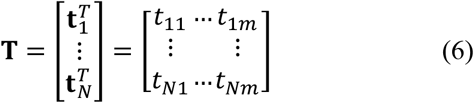

For these linear equations, an optimal solution was proposed in [12] as:

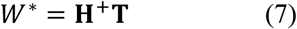

where: **H**^**+**^: Moore-Penrose generalized inverse function (+).

The optimal solution is then used in Eq. (7) to issue prediction for any given input vector **x**.

### 2.3. Model development

A three-layered architecture was adopted for ELM model development (Fig. 2). The first layer (input layer) used the data (Table 2) as inputs. The output layer had one neuron representing the New COVID-19 Cases (NCC). For the single hidden layer, seven neurons a maximum were tested for each model.

**Table 2:** Performances parameters of ELM models with various number of neurons in hidden layer (the bold refers to the best model)

| ELM | $R_{TRAIN}$ | $R_{TEST}$ | $All R$ | $MSE$ | $RMSE$ | $MAE$ | $NSE$ | $IO$ |
| --- | --- | --- | --- | --- | --- | --- | --- | --- |
| 3-2-1 | 0.8481 | 0.7944 | 0.8876 | 511.91 | 22.6256 | 17.2148 | 0.7879 | 0.8329 |
| 3-3-1 | 0.9984 | 0.9988 | 0.9985 | 6.83 | 2.6145 | 1.9017 | 0.9971 | 0.9915 |
| 3-4-1 | 0.9999 | 0.9973 | 0.9998 | 0.80 | 0.8980 | 0.6117 | 0.9996 | 0.9974 |
| **3-5-1** | **0.9999** | **0.9998** | **0.9999** | **0.16** | **0.4114** | 0.2912 | **0.9999** | **0.9988** |
| 3-6-1 | 0.9999 | 0.9979 | 0.9998 | 0.66 | 0.8137 | 0.5102 | 0.9997 | 0.9979 |
| 3-7-1 | 0.9999 | 0.9991 | 0.9999 | 0.25 | 0.5002 | **0.2638** | 0.9998 | 0.9988 |

**Figure 1:**
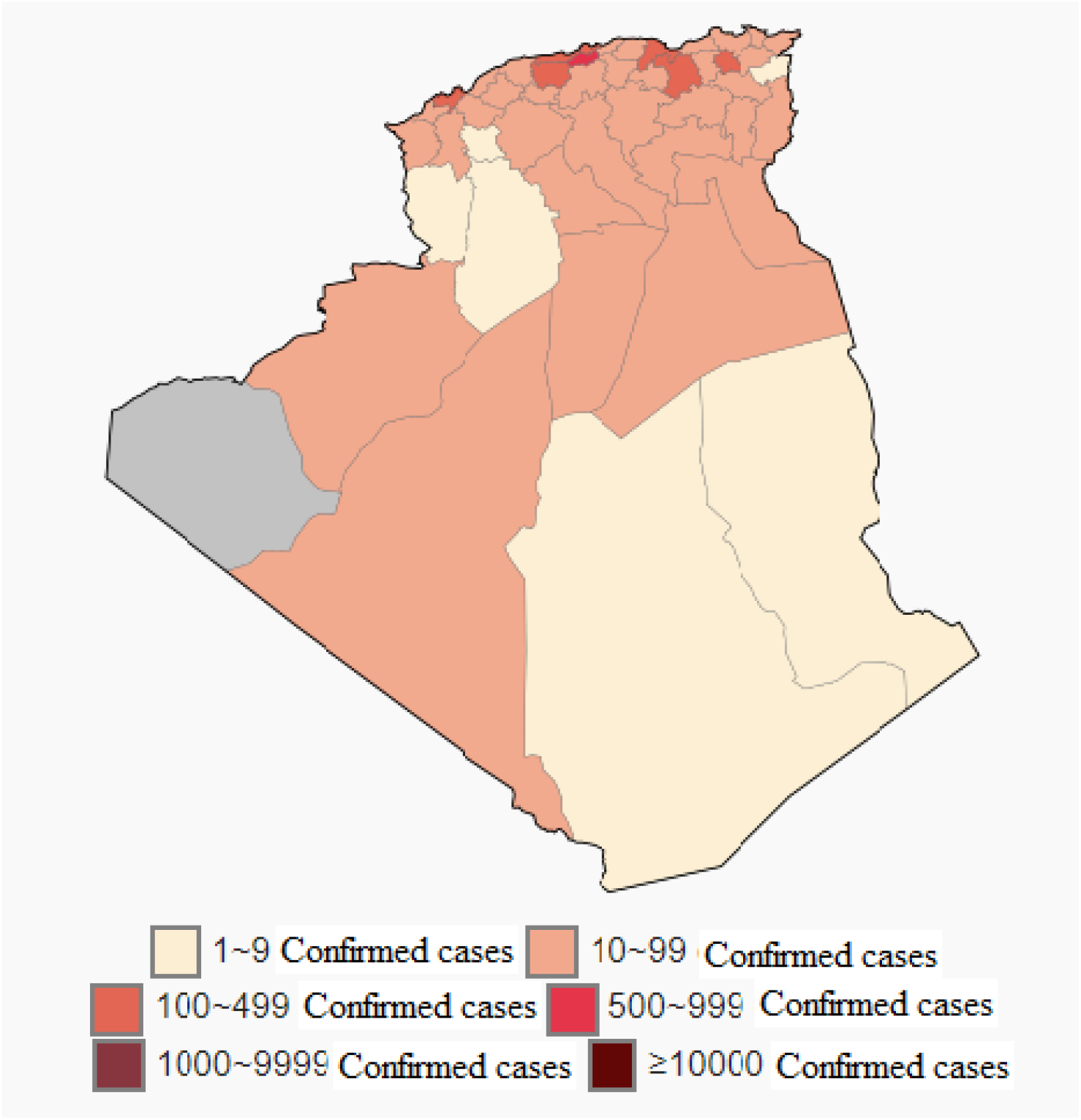
Map of the Covid-19 pandemic in Algeria [18]

**Figure 2.**
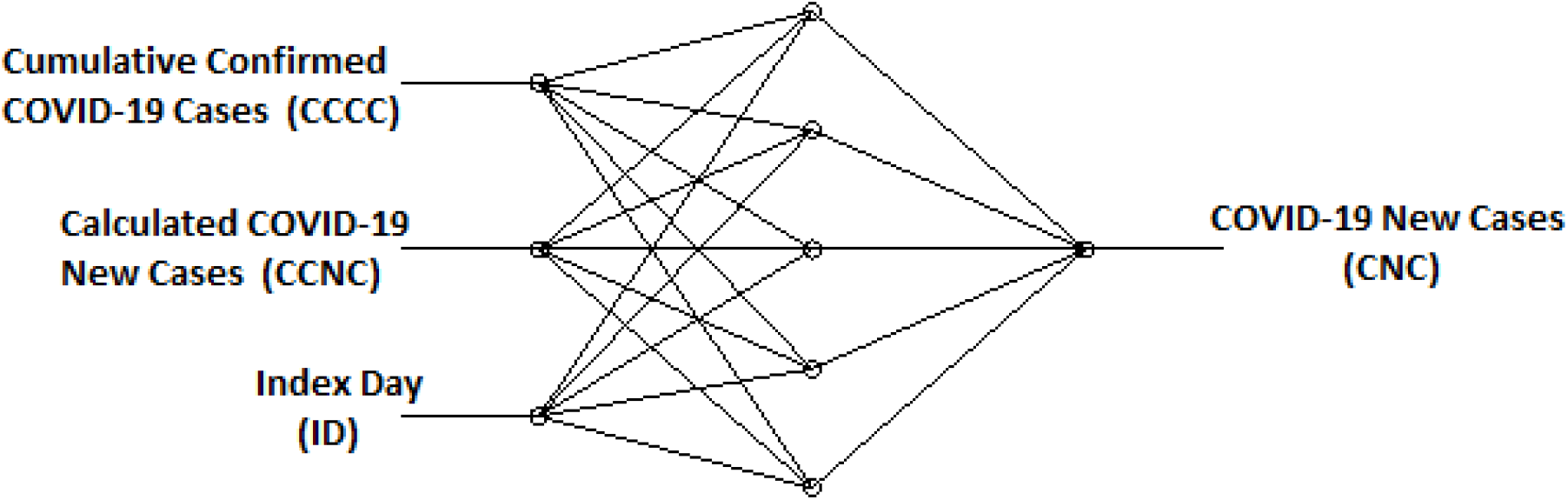
Architecture of the proposed ELM model (3-5-1).

Finding the appropriate number of nodes in the single layer hidden in the extreme learning machine neural network is a very difficult and critical task. A model with a large number of hidden nodes will have an excessive configuration, will follow the noise in the data and consequently will require more time in training phase, with the risk that overfitting occurs. The generalization properties during the test phase can be considerably influenced [13].

Some general rules for selecting the number of nodes in hidden layer (NNHL) are presented below:

1. Hecht-Nielsen [20] used the Kolmogorov theorem to prove that:

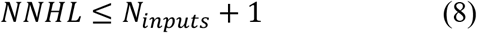
2. Hecht-Nielsen [21] suggest that it should be:

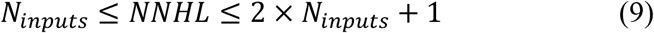
3. Masters [22] suggests that the ANN architecture should resemble a pyramid with:

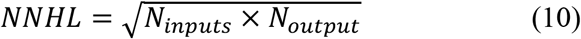

Where:

*N*_*inputs*_ and *N*_*output*_ are respectively, the numbers of inputs and output.

For determining the optimum number of neurons in the hidden layer, initially, two neurons were tested. Subsequently, the number of neurons was gradually increased to 7 by an interval of one. Sigmoid activation function (*sig*) was adapted for all the ELM models tested in the hidden layer and linear function as transfer function (*pure line*) in the output layer.

### 2.4 Criteria for evaluation

The best ELM configuration is adopted based on the minimum statistical error measurements (*MSE, RMSE, MAE*), the highest values of the performance parameters (*R*^*2*^, *NSE, IO*) between the calculated and predicted values using six statistical measures:

1. Mean-square error (*MSE*);
2. Root-mean-square error (*RMSE*);
3. Mean Absolute Error (*MAE*)
4. Correlation Coefficient (*R*);
5. Nash–Sutcliffe Coefficient of efficiency (*NSE*);
6. The overall index of model performance (*OI*).

These parameters were calculated to check the performance of the developed models. These statistical performance evaluation criteria are expressed as follows:

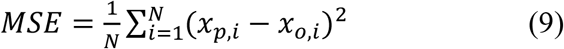

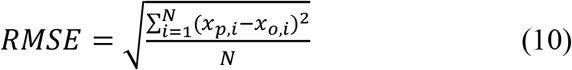

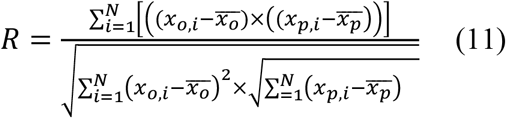

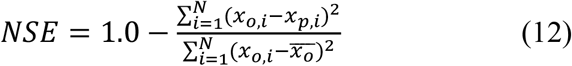

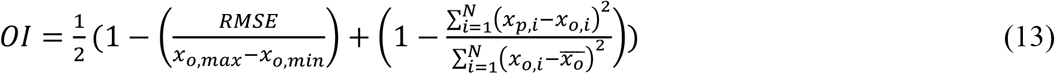

Where: *x*_*o,i*_ = observed value; *x*_*p*,i_ = predicted value; *N* = number of observations; *x*_*o,max*_ = maximum observed value; *x*_*o,min*_ = minimum observed value; and 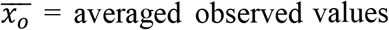; 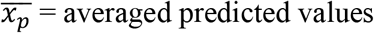.

Legates and McCabe [23] mentioned that *RMSE* has the advantage of expressing the error in the same units as the variable, so, providing more information about the efficiency of the model. A model with a lower RMSE value will have a high accuracy prediction. In addition a model with high value of *OI* means a perfect fit between the observed and predicted data [24].

## 3 RESULTS AND DISCUSSION

The ELM models development and construction process consists of the following five steps:

1. Selection of the input and the output data for training ELM model (data set).
2. Normalization of the input and the output data attributes.
3. Training of the normalized data using a hybrid-learning algorithm.
4. Testing the goodness of fit of the model.
5. Comparing the predicted output with the target output.

The input and the output data was normalized using min-max normalization; the data are normalized to values between 0 and 1 using the equation:

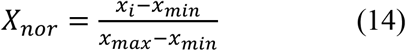

Where *x*_*min*_ and *x*_*max*_ are the minimum and maximum values of *x*_*i*_, respectively.

After obtaining the normalized data, the next step is to train the input data using the proposed ELM model. ELM model uses Gradient-based Algorithms with Single Hidden Layer Network. The algorithm, by default, takes 80% of the input data for training. So out of 56 data, 45 data was randomly selected from the normalized dataset for training the model. The rest, 11 data, are kept for testing the model. After the testing is done, the ELM model is saved. Mean-squared error (*MSE*), Root Mean-Squared Error (*RMSE*), and Mean Absolute Error (*MAE*) between calculated and predicted values are used as a performance index of the model accuracy, as it is shown in Table 2.

The trials started using two neurons/nodes in the hidden layer as the initial start. As shown in the first row in Table 2, the values of *MSE* for CNC was 511.91.

The values of *RMSE*, respectively, for the first model (3-2-1), was 22.62. *MAE* values for CNC were 17.21 for the ELM model with two neurons in the hidden layer.

The value of *R* in the training phase was estimated at 0.8481, which shows a good similarity between the values of calculated and predicted CNC during the training process. For the testing phase, the value of *R* was 0.7944. The testing step reveals that the ELM model generalization ability is very satisfactory.

Increasing the neurons in the hidden layer gave a clear and marked improvement in the ELM model performance. Arriving at the topology 3-5-1, the lowest values of *MSE* are obtained, 0.16 for CNC. For this same topology, the lowest values of *RMSE* and *MAE* are obtained.

The highest value of *OI* was respectively, 0.9988 is obtained with the ELM model 3-5-1. The analysis of the coefficient of efficiency (*NSE*) of the ELM models shows that the 3-5-1 model has the best value, namely 0.9999.

It can be noted from the fig. 3 that predicted CNC values are quite close to the calculated CNC values, as their coefficient of determination (*R*^*2*^) value are very close to unity.

**Fig. 3:**
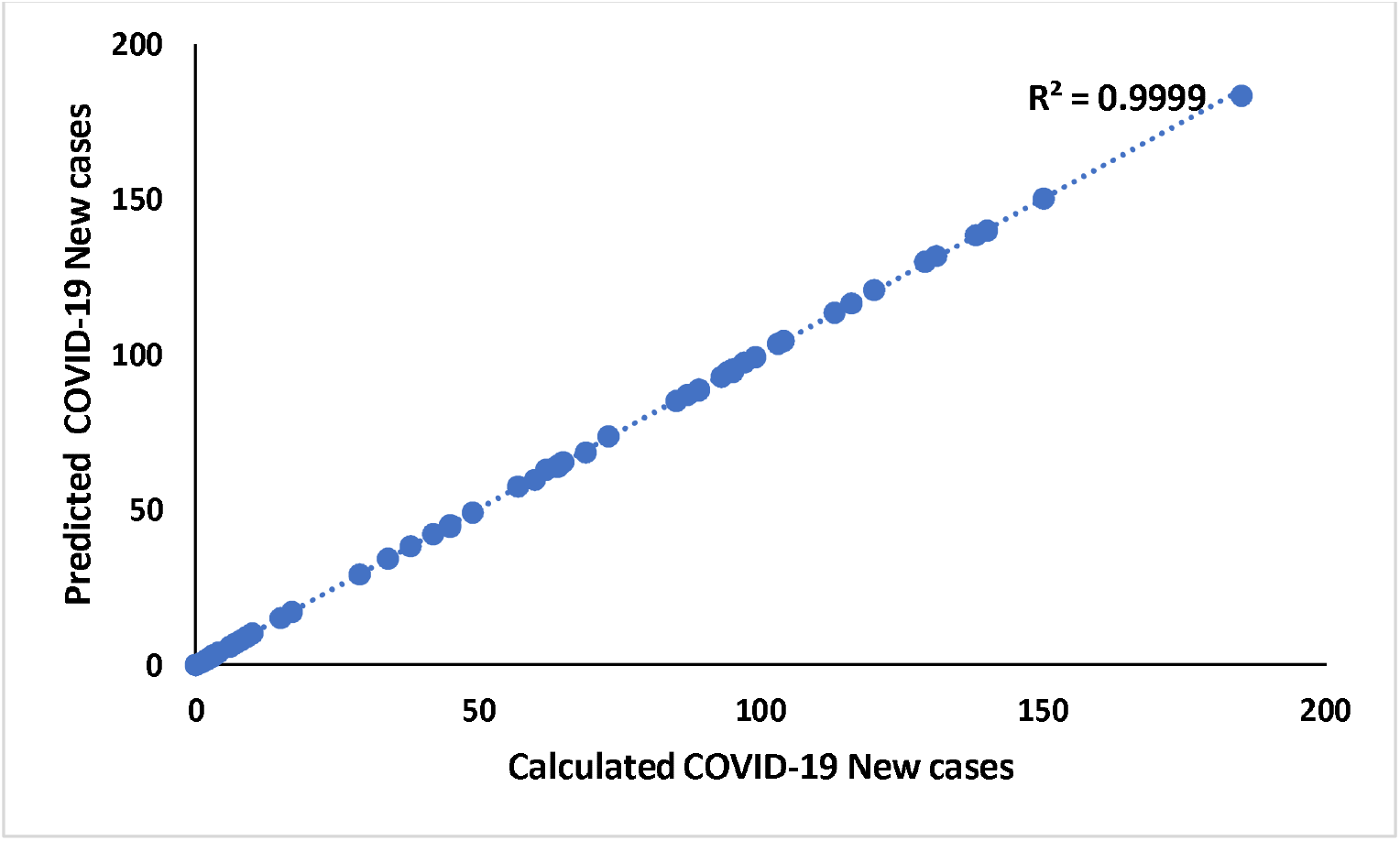
Calculated COVID-19 new cases vs predicted new COVID-19 cases.

As a result, the most appropriate ELM architecture is 3–5–1 (bolded in Table 2), as this gave the best CNC prediction with the lowest error (*MSE, RMSE*, and *MAE*; and the maximum *R*_*all*_). The best architecture of the developed ELM model is schematically illustrated in Fig. 2.

## 4 CONCLUSION

This paper proposed an extreme learning machine model (ELM) as machine learning technique for a prediction task. The proposed ELM model has a high ability to predict the number of new COVID-19 cases in Algeria with high accuracy, lowest errors (in terms of *MSE, RMSE*, and *MAE*), and highest performances parameters (*NSE, IO*, and *R*^*2*^). According to the precise results obtained by the proposed ELM model, it can be used as a decision support tool to manage public health medical efforts and facilities against the COVID-19 pandemic crisis.

## Data Availability

All the data used in this research was was donwloaded from the web site:
https://en.wikipedia.org/wiki/COVID-19_pandemic_in_Algeria

